# Complicated Common Variable Immunodeficiency is Driven by Aberrant IL-10/IL-21 Signaling and Predisposed Polygenic Risk

**DOI:** 10.1101/2022.06.20.22276681

**Authors:** Humza A. Khan, Utkucan Acar, Alexis V. Stephens, Timothy J. Thauland, Manish J. Butte

## Abstract

**Background:** The inborn errors of immunity (IEI) that include defective antibody responses are clinically heterogenous, especially the common variable immunodeficiency (CVID) phenotype that includes low immunoglobulin levels and impaired humoral responses to antigens. Beyond recurrent infections, many with the CVID phenotype develop non-infectious complications (NICs), including autoimmunity and lymphoproliferation, that confer a high rate of morbidity and mortality. At present, it is unknown what genetic and functional factors predispose patients to NICs.

**Objective:** We aimed to discover the pathobiology underlying complicated CVID (CVIDc).

**Methods:** In a heterogenous group of 12 CVIDc patients, we conducted whole exome sequencing and high-throughput signaling assays by multiplexed phospho-mass cytometry. The immune deficiency and dysregulation activity (IDDA) score was used to determine the burden of NICs in individual patients. We integrated polygenic risk scores to determine the role of common background variants in the pathogenesis of CVIDc.

**Results:** In CVID patients with high IDDA scores, there was aberrant increased phosphorylation of STAT1 and STAT3 upon stimulation with IL-10 or IL-21. Furthermore, common variants related to high eosinophil count and allergy/eczema confer a higher likelihood of autoimmunity in CVID.

**Conclusion:** Variants in loci related to high eosinophil count/function and over-reactive IL-10 signaling are associated with the development of autoimmune disease and NICs in CVID.

**Clinical implications:** It may be possible to manage CVIDc through modulating IL-10 and IL-21 signaling pathways. Polygenic risk scoring may predict the development of autoimmune complications in CVID patients.

## Introduction

The phenotype of Common Variable Immunodeficiency (CVID) is among the most common inborn error of immunity (IEI), affecting around 1 in 25,000 people(1,2). This phenotype is recognized by laboratory findings of low immunoglobulin levels and defective antibody responses plus a relevant clinical history. Most patients with the CVID phenotype endorse a history of prolonged or recurrent sinopulmonary infections. On the other hand, there is considerable variability in their non-infectious complications (NICs)(3): inflammatory and autoimmune complications affecting the lungs, GI tract, hematopoietic cells, joints, and other systems. It is estimated that 30-50% of CVID patients present with autoimmune and inflammatory features in addition to recurrent infections(1,2,4). It is these patients with NICs (also called complicated CVID, CVIDc) who face a significantly higher mortality rate than their counterparts who suffer only infectious complications(1). If we could predict early which CVID patients are likely to develop NICs we could treating these complications before they cause lasting harm. Thus, there is an urgent need to better understand NICs in CVID.

The etiology of NICs is thus far unknown, but numerous efforts have sought to link NICs to an underlying genetic aberration causing the CVID phenotype. Indeed, many monogenic variants have been identified that explain both immune dysregulation and also a defect in B-cell development, maturation, signaling, and survival. Pathogenic variants in genes related to B-cell signaling (e.g., *NFKB1, NFKB2, PIK3CD, CD20*) are known to be causes of CVID, but less than half the CVID patients carrying pathogenic variants in these genes have NICs, suggesting that monogenetic variants are not sufficient to explain the CVIDc phenotype. Moreover, over half the CVID cases do not have a attributable monogenic cause(5–7). Copy number variation, somatic mosaicism, and other factors highlight why genetic diagnoses in CVID are difficult(8,9). Finally, non-genetic features like changes in the gut microbiome offer correlations in those with NICs(10). For these reasons, approaches to understand the tendency to NICs have employed immune phenotypes or functional studies. Previous studies have identified dimished switched memory B cells, increased levels of inflammatory cytokines, global transcriptional response to LPS, and monocyte activation in patients with CVIDc(11–15). These findings have provided some insight into the pathogenesis of CVIDc, suggesting a mechanism of overactive signaling in monocytes, or excessive T-cell infiltration into otherwise healthy tissues. Our previous work identified functional defects in signaling pathways in IEIs using CyTOF(16). But whether CyTOF or other approaches can identify subjects with NICs has not been explored.

Disturbances in B-cell development pathways via multiple polygenic variants can also cause CVID(17). The phenotypic presentation of CVID and especially CVIDc may thus best be understood from a polygenic and epistatic standpoint. Distinct from the paradigm of immune diseases that arise from rare, monogenic defects, it is known that common variants in multiple genes influence the presentation of more complex traits. Polygenic risk scores (PRS) are individual-level scores calculated from the presence of variants correlated to traits via genome-wide association studies (GWAS). PRSs have proven their potential use to stratify risks in patients in various conditions (such as schizophrenia and atherosclerosis)(18,19). The same methodology could very well apply to complicated versus uncomplicated CVID. We sought here to understand how the architecture of *common* genetic variants influences the CVIDc phenotype, and to understand the contribution of aberrant variants in signaling pathways with empirically-identified defects in signaling pathways.

## Methods

### Subjects

All subjects provided informed consent for a research protocol approved by the Institutional Review Board of the University of California Los Angeles.

### Stimulation Phospho-Mass cytometry

Heparinized blood samples were taken from CVID patients and age- and biological sex-matched controls.

In our studies, we sought to include the myeloid cells that might otherwise be removed in traditional gradient separation of PBMCs. We thus started our work with whole blood(20). However, to prevent serum IgM from competing with naïve B-cell-surface IgM for binding to anti-IgM antibodies, samples were first washed in PBS+1% bovine serum albumin (BSA). To prevent nonspecific binding of antibodies to Fc receptors, samples were then supplemented with 2 mM CaCl□ and incubated with FcX (BioLegend, 422302) according to manufacturer instructions. Samples were then stained with surface antibodies for 30 minutes at room temperature.

After incubation, samples were washed and then aliquotted for stimulation with a variety of individual stimuli: IL-2 (10 ng/mL, Peprotech, 200-02), IL-6 (10 ng/mL, Peprotech, 200-06), IL-10 (10 ng/mL, Peprotech, 200-10), IL-21 (50 ng/mL, Peprotech, 200-21), IFN-α (50 ng/mL, Cell Signaling Technology, 8927SC), IFN-γ (10 ng/mL, Peprotech, 300-02), R848 (1 μg/mL, Invivogen, vac-r848), PAM3CSK4 (5 μg/mL, Invivogen, tlrl-pms), LPS (0.5 μg/mL, Sigma-Aldrich, L4391), PMA/Ionomycin (10 ng/mL/1 μg/mL,Sigma-Aldrich, P1585, I3909)) or PBS as control at 37 °C for 15 minutes.

Stimulation was arrested and red blood cells were lysed together by using Lyse/Fix Buffer (BD Biosciences, 558049). Samples were then washed and permeabilized with methanol. After permeabilization, samples were stained for intracellular targets and incubated with Fix/Perm Buffer (Fluidigm, 201067) and 125 nM Iridium DNA intercalator (Fluidigm, 201192B). Samples were finally washed and then run on a Helios mass cytometer.

Antibodies used in the assay are available in supplementary **Table S2**.

### Gating

The CytoExploreR package in R was utilized for gating (21). The gating scheme is shown (**Fig. S3**). Manual gates were drawn with the same cellular populations analyzed from date to date.

### Gaussian Mixture Modeling

Bimodal phosphorylation responses were modeled using a 2-component Gaussian mixture modeling (GMM) framework using the expectation-maximization algorithm(22). GMMs were initialized with 2-component k-means clustering of phosphorylation responses. Up to 1000 iterations were performed on the data or if the difference between the log-likelihoods of two iterations reached less than 10^6. Models were constrained such that if two components were within 1 unit of each other or if one component represented less than 5% of the data, it was replaced by the larger model. Variance was also restricted to greater than .01. Values above 0 were considered for model fitting. Examples are included in **Fig. S4**.

### Batch Effect Correction

Phosphorylation values from different experimental dates were adjusted to allow for accurate comparisons despite batch effects. The unstimulated Gaussian mixture model phosphorylation values from a date’s control were used to adjust each cell type-signaling pathway combination for all stimuli as was previously described(16).

Values of zero were removed from the data and are common statistical noise in all CyTOF datasets.

### Exome sequencing

DNA from whole blood was extracted and then sent to Psomagen for whole-exome sequencing using the Agilent capture kit.

### Polygenic Risk Score Calculations

VCFs of patients with CVID from the European Genome-Phenome Archive were processed into CSVs, then additively joined to recently described polygenic risk scoring files(23).

### Gene ontology analysis

SnpXplorer was used to analyze SNP-sets for gene ontology, annotation, and relation to GWAS traits(24).

### Statistical analysis

In all applicable comparisons, bootstrapping was used to generate reference distributions of given statistics. This mitigates the contribution of outliers to the generation of a statistic and allows for the calculation of 95% confidence intervals. Significance testing was done using permutation testing.

### Code Availability

Scripts are hosted at https://github.com/humzalikhan/CVID_CyTOF_PRS or otherwise are available upon request.

## Results

### Patient cohort and disease scoring

We performed Cytometry by Time of Flight (CyTOF/mass cytometry) on whole blood on 12 patients with CVID (**Fig. 1**).

**Fig. 1.**
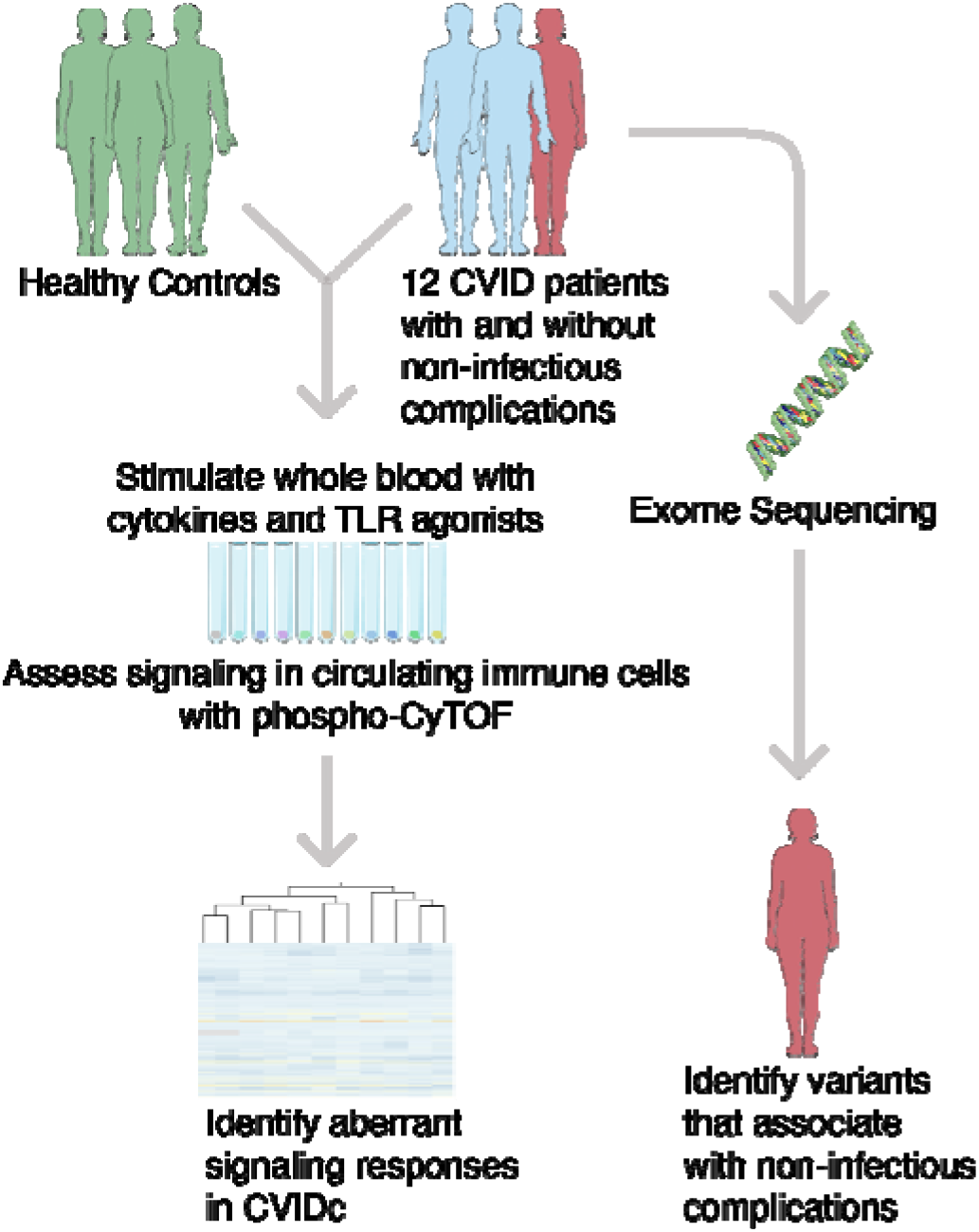
Schematic of study design Whole exome sequencing was also performed. Our cohort represented a diverse group, with 5 males and 7 females with an average age of 44 years (**Fig. 2a**). Only one patient had a known pathogenic variant in genes associated with monogenic cause for CVID (*NFKB1* haploinsufficiency); others had variants of unknown significance in other genes associated with CVID such as *AIRE, MALT1*, and *RASGRP3* (data not shown).

Given the heterogenous presentation of immune dysregulation in CVID, we used the recently described immune deficiency and dysregulation activity (IDDA) score to assess the extent of a patient’s non-infectious complications within our cohort (25,26). Patients above an IDDA score of 12 were considered IDDAhi, and others were considered IDDAlo. Patients who were classified as IDDAhi had significantly higher incidence of autoimmune cytopenias, enteropathies, lymphoproliferation, and lung disease (**Fig. S1**). Of note, our cohort did contain one outlier patient, with especially high disease burden and high IDDA score. To avoid excessive value to this one individual in downstream analyses, we employed bootstrapping/permutation-based analyses. Overall, we have a varied patient population composed of individuals with and without CVIDc.

### Cellular subsets correlated with disease burden in CVID

We performed resampled correlation analysis between cell frequencies in patients and IDDA scores. We found that patients with higher frequencies of CD21lo B cells among all B cells were highly correlated to higher disease burden (**Fig. 2b**). CD21lo B cells are transitional B cells that are autoreactive and contribute to autoimmunity (27–30). This finding supports previous work demonstrating the link between CD21lo B cells and CVIDc (15). Furthermore, the frequencies of all subsets of monocytes (CD16lo, mid, and hi) were positively associated with higher disease activity. We view high monocyte frequencies as a proxy for monocyte activation, as monocyte activation induces effector differentiation and proliferation (31). Indeed, monocyte activation and proliferation has been related to NICs in CVID (13,14). Higher proportions of switched memory B cells (CD27+ IgD-IgM-), regulatory T cells, and naïve T cells were associated with lower IDDA scores (**Fig. 2c-d**). These findings support that low IDDA scores may be driven by the presence of switched memory B cells, which have undergone receptor editing and lack polyreactivity, or regulatory T cells, which also oppose autoimmunity.

**Fig. 2.**
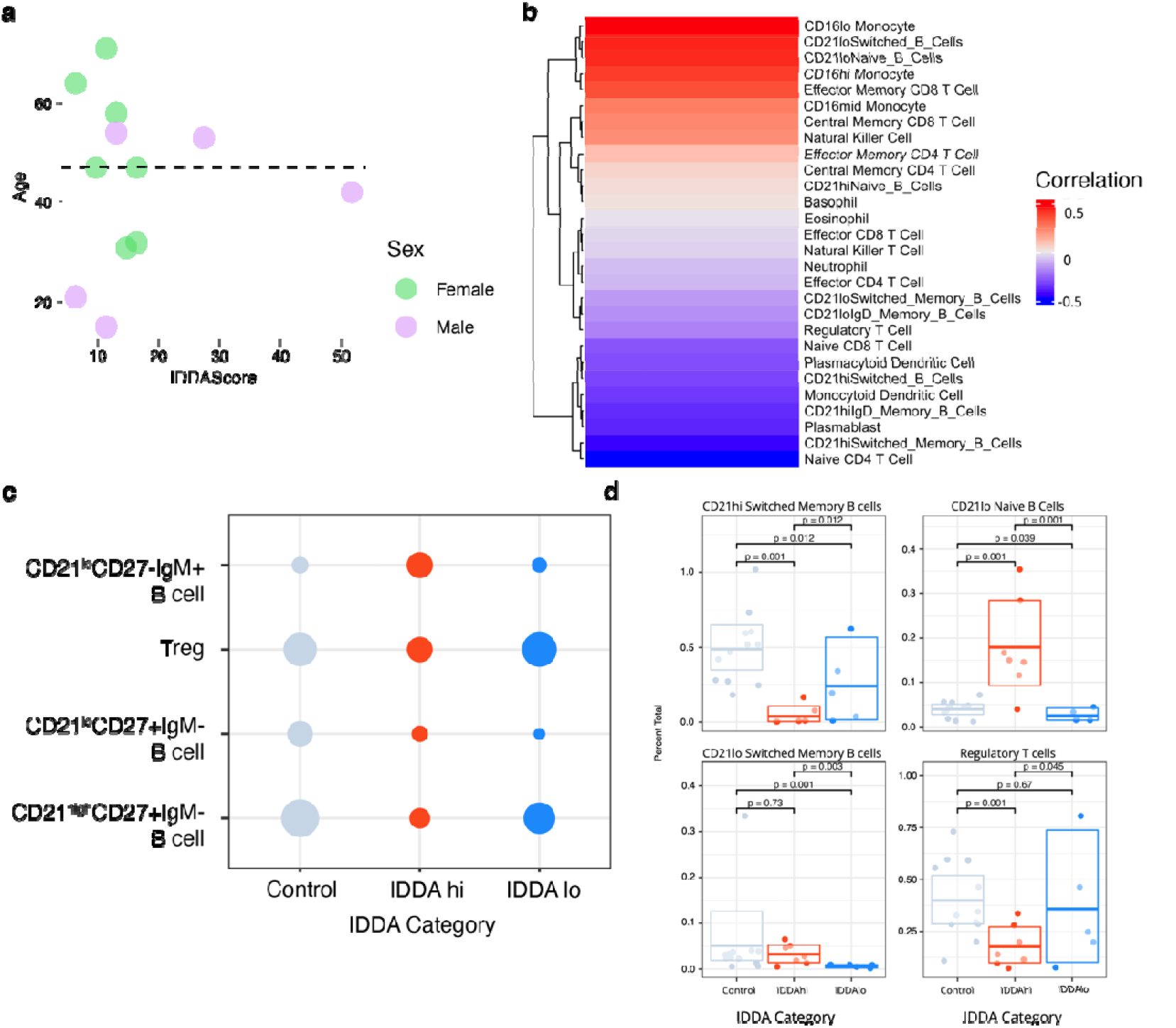
Cohort description and cellular frequencies associated with disease course in CVID. **a)** distribution of age, sex and Immune deficiency and dysregulation activity of CVID cohort. **b**. Correlation analysis of cell type frequencies and IDDA scores. **c**. Cell subtypes with differential abundance between IDDAhi and IDDAlo patients. Circle sizes are relative and relate the proportions of each cell type among peripheral blood lymphocytes. **d**. Proportions of each identified cell type in c.

We compared frequencies of immune cells between healthy controls, IDDAhi patients, and IDDAlo patients. IDDAhi patients had much lower percentage of switched memory B cells and higher percentage of CD21lo B cells in comparison with both healthy controls and IDDAlo patients. Additionally, FOXP3+ regulatory T cells were diminished in IDDAhi patients (**Fig. 2c-d**). These findings are in line with the previous observations and support the hypothesis that autoimmunity in CVIDc develops via lack of proper immune regulation by regulatory T cells and an expansion of the CD21lo immature B cell subset.

### Overresponsive IL-10/IL-21 Signaling found in CVIDc

Since overresponsive cellular signaling has previously been linked to autoimmunity, we investigated cellular signaling in CVIDc using a multiplexed stimulation assay using phospho-CyTOF (32).

We employed heatmap analysis of these results to visualize the separation between healthy controls and IDDA-stratified CVID patients. We find a clear distribution of these responses, with healthy controls and IDDAlo patients barely responding to IL-10 in the STAT1/STAT3 pathways, while IDDAhi patients signaled much more strongly than either of the other subgroups (**Fig. 3a**). Similar results were found for IL-21 and STAT1/STAT3 (**Fig. 3b**).

**Fig. 3.**
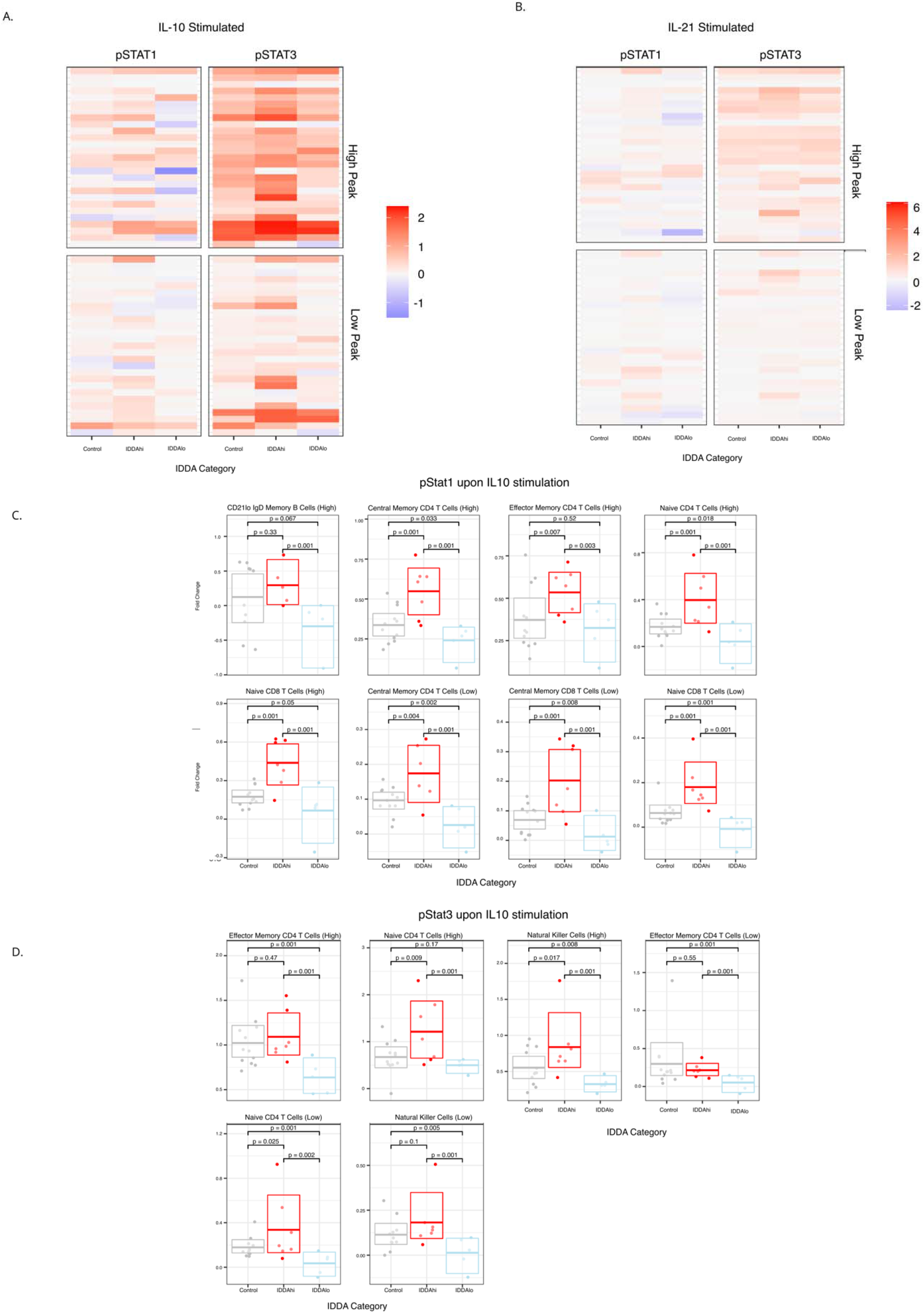
Profound signaling defects in the IL-10 and IL-21 axes are correlated non-infectious complications in CVID. A. Heatmap indicating aberrant IL-10 driven pSTAT1 and pSTAT3 signaling. B. Heatmap indicating aberrant IL-21 driven pSTAT1 and pSTAT3 signaling. C. T cell subsets associate with aberrant IL-10 signaling in CVIDc.

Furthermore, IL-10 stimulation induced strong decreases in phosphorylation of AKT, CREB, and S6 in healthy controls and IDDAlo patients. However, in IDDAhi patients had increased levels of phosphorylated AKT, CREB, and S6 in response to IL-10 (**Fig. S5**).

We then performed hypothesis testing of which cell-type subsets displayed aberrant signaling in CVIDc. After stimulating cells with IL-10, we found that patients with CVIDc (IDDAhi patients) had increased responses in the phosphorylation of STAT1 and STAT3 in many T cell subsets and natural killer cells (**Fig. 3c**). IL-21 also induced similar over-responsivity of STAT1 and STAT3 phosphorylation in these cell types (**Fig. 3b, Fig. S5**).

An opposite effect was found with the phosphorylation of S6, while also including CD16hi/lo monocytes, monocytoid dendritic cells, and neutrophils (**Fig. S5**).

### Eosinophil and Allergy/Eczema Polygenic Risk Scores Confer Higher Likelihood of Developing Autoimmune NICs in CVID

To expand the findings in our small local cohort of patients, we utilized genome sequencing data of IEI patients from the National Institute for Health and Care Research (NIHR) Bioresource. This cohort included 451 patients with CVID and phenotyping data on their comorbidities (380 with full phenotyping on autoimmunity). This large group represents around 1/3 of all CVID patients registered in the UK(33).

We explored polygenic risk scores (PRS) to find the effect of common variants related to some traits. We performed odds-ratio analysis, focusing on PRSs where patients in the 9^th^ and 10^th^ deciles for a given PRS had high odds-ratios for autoimmune complications in CVID. We chose to examine autoimmune complications specifically given that this is a particularly discriminating feature between CVID and CVIDc. We found two PRSs that satisfied our decile criterion: one for eosinophil count and one for diagnosed allergy/eczema (**Fig. 4a, Fig. 4b**).

**Fig. 4.**
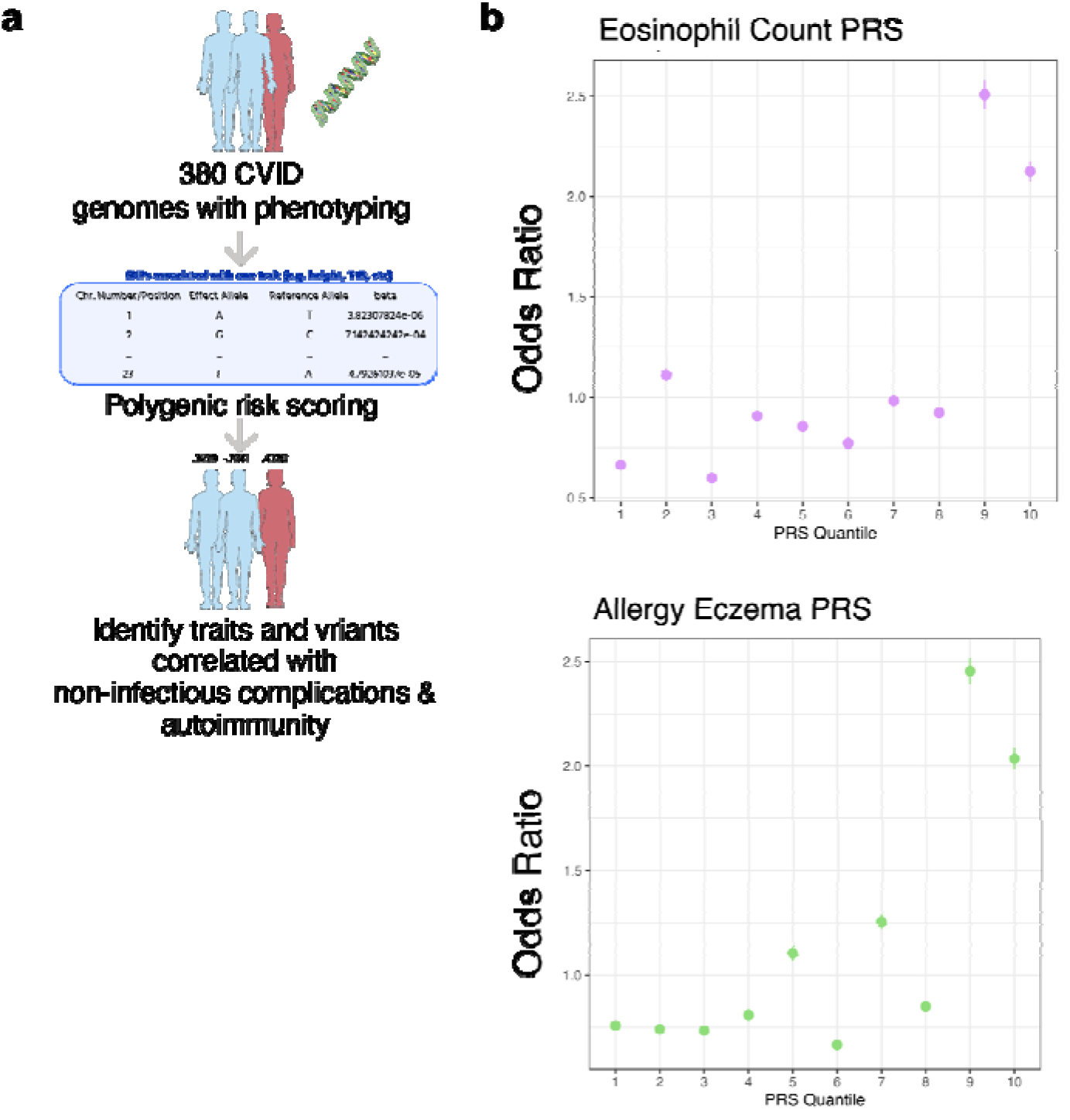
Eosinophil count and allergy/eczema polygenic risk scores associate with complicated CVID. A. Schematic of computational analysis of NIHR CVID cohort. B. CVID patients with autoimmunity are enriched in the top two deciles of eosinophil count/allergy and eczema polygenic risk scores.

We then performed gene ontology and genome-wide association study (GWAS) catalogue analysis on the 10,000 most contributing SNPs of the two relevant PRS variant sets. The two variant sets are not orthogonal and contain some of the same SNPs. This is seen with many genes sharing the same ontology. We found that SNPs from both PRSs related to the GWAS catalogue traits of type II diabetes, hemoglobin measurements, and lymphocyte count indicating a strong inflammatory signal in CVIDc.

## Discussion

To our knowledge, no other work has utilized both genomics and high-dimensional phospho-proteomics to explain the differential pathogenesis of an inborn error of immunity. We found signaling defects in patients with complicated CVID, with the potential to use these as a predictive measure of autoimmunity and other complications in within CVID. Furthermore, we found polygenic variants influencing the likelihood of a patient’s CVID to become complicated with extra-infectious symptoms. This is likely the first application of polygenic risk scores to the stratification of a rare immune disease.

IL-10 shows pleiotropic effects, often noted for suppressing the inflammatory response. However, higher IL-10 production has been linked to autoimmune disease and other immune regulatory disorders (34,35). Additionally, patients with CVIDc were found to have higher levels of IL-10 circulating compared to healthy controls and other antibody deficiencies(14). Our results showing enhanced STAT1/3 and AKT pathway phosphorylation in response to IL-10 presents a new paradigm for understanding complications in CVID: perhaps a non-canonical IL-10 positive feedback loop erroneously increases pro-inflammatory signaling, which leads to the production of IL-10, a quintessentially anti-inflammatory cytokine.

Furthermore, IL-21 has been shown to be important for the Th17 axis of autoimmunity(36). It is possible that IL-21 is driving an alternative route of autoimmunity in CVID or synergizing to drive non-infectious complications in CVID.

Polygenic risk has previously been noted in small-scale studies of CVID(17). By extending this on a much larger scale, we have found variant sets that may predispose CVID patients to complications. Eosinophilia and allergy/eczema can be associated with CVID, though are not necessarily thought of as major complications of the disease. IL-10 has been shown to be an overexpressed cytokine in atopic dermatitis, related to Th2 skewing in this disease(37). Our finding may provide a link between CVID, allergy/eczema pathways, and Th2/Th17-driven autoimmunity. Interestingly, while eosinophil count did not associate with IDDA score, SNPs modulating eosinophil function likely augment Th2 immunity and could cause NICs through this axis (**Fig. 1C**).

Continued experiments are necessary to understand the role of IL-10/IL-21 biology in CVID; potentially, future biologics blocking IL-10/IL-21 or their receptors will prove to mitigate inflammatory, non-infectious complications of CVID. Furthermore, polygenic risk scoring from diverse datasets may also contribute to identifying CVID patients at risk for complications and allow them to get treated promptly.

## Data Availability

All data produced in the present work are contained in the manuscript, hosted at https://github.com/humzalikhan/CVID_CyTOF_PRS, or otherwise are available upon request.

## Funding Acknowledgments

This study was supported by the NIH (R01AI153827 to MJB) and a gift from the Jeffrey Modell Foundation.

## Abbreviations

CVID, CVIDc, NICs, CyTOF, IEI, IL-10

## Acknowledgements

We thank Dr. Alejandro Garcia and Miriam Guemes of the UCLA Jonsson Comprehensive Cancer Center (JCCC) and Center for AIDS Research (CFAR) Flow Cytometry Core Facility for running samples. The UCLA JCCC Flow Cytometry Core is supported by NIH P30 CA016042 and 5P30 AI028697.

We gratefully acknowledge early advice on our CyTOF panel and protocols offered by Drs. Holden Maecker (Stanford), William O’Gorman (Genentech), and Elena Hsieh (University of Colorado). We acknowledge Rohit Srinivas for graphic design help with figures. We thank members of the Butte laboratory for feedback on the manuscript.

This work used computational and storage services associated with the Hoffman2 Shared Cluster provided by UCLA Institute for Digital Research and Education’s Research Technology Group.

## Notes

### Competing Interest Statement

The authors have declared no competing interest.

### Funding Statement

This study was supported by the Jeffrey Modell Foundation through a grant from Takeda; NIH/NIAID R01AI153827 to MJB

### Author Declarations

IRB of the University of California Los Angeles gave ethical approval for this work.

