## Supplemental Figures for "Complicated Common Variable Immunodeficiency is Driven by Aberrant IL-10/IL-21 Signaling and Predisposed Polygenic Risk"

**Fig. S1.** IDDA breakdown of cohort by associated conditions.

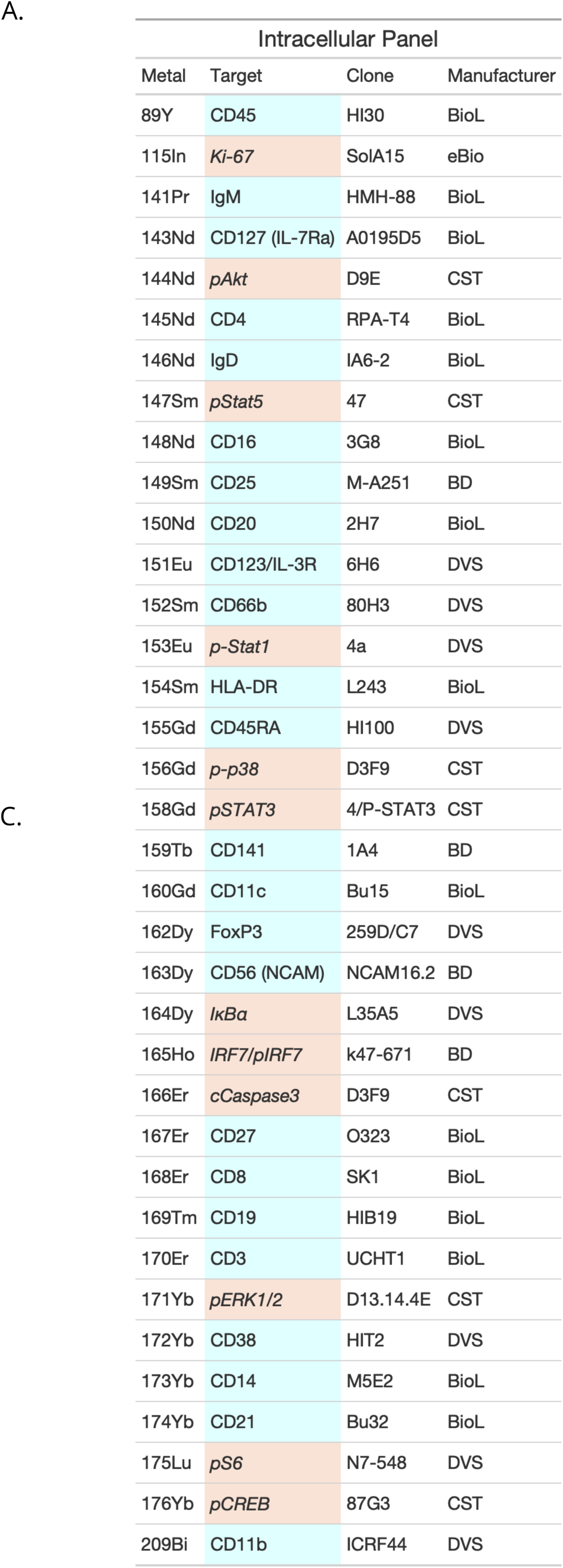

**Fig. S2**. Mass cytometry antibody panel.

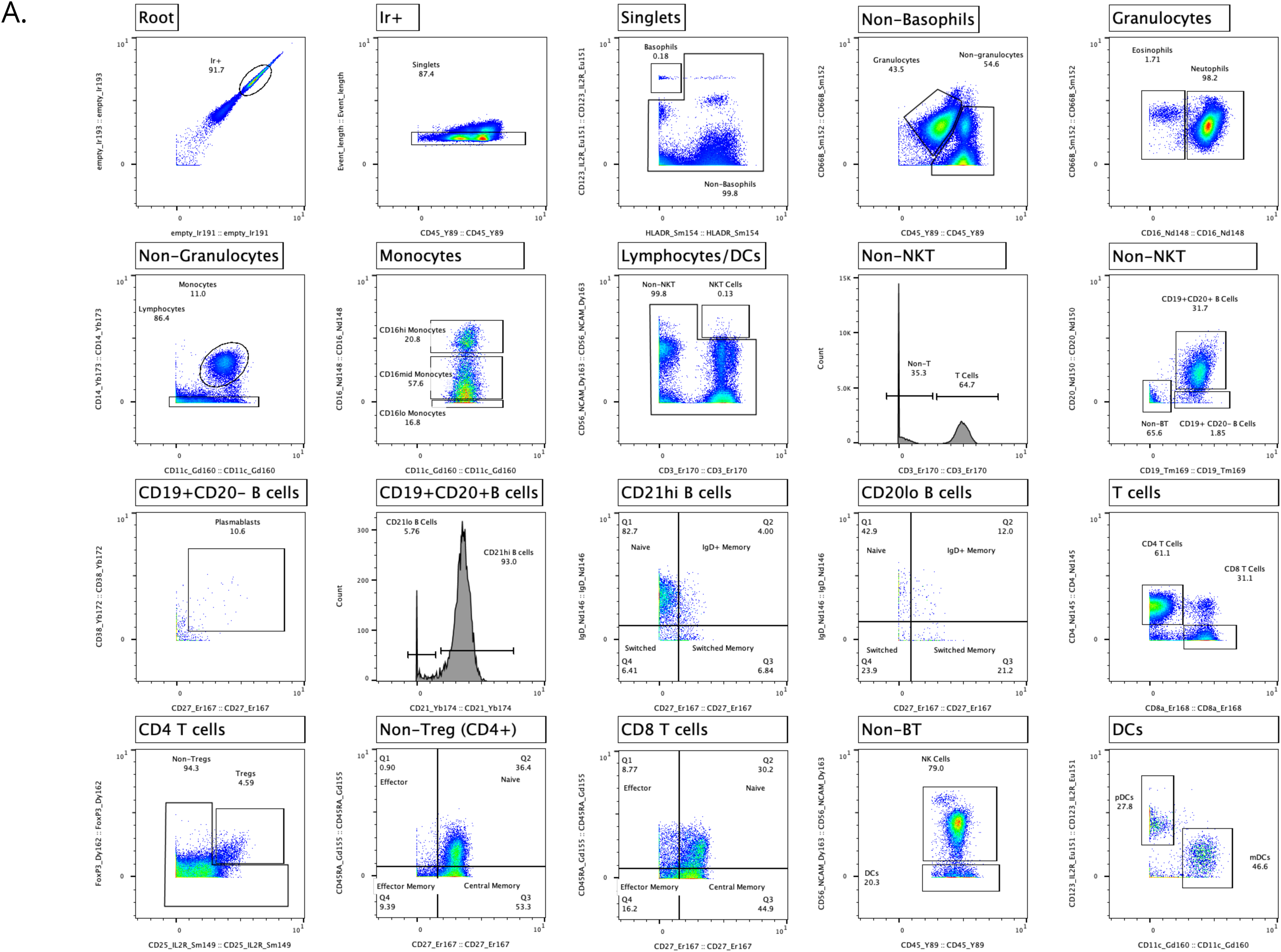

**Fig. S3.** Gating scheme for phospho-mass cytometry.

**Fig. S4**. Example of Gaussian Mixture Model (GMM) approach to analyzing cellular signaling.

**Fig. S5**. IL-21 - pSTAT1/pSTAT3 and IL-10 pS6, pCREB, and pAKT axes associate with CVIDc.

Figure S6. A. SNP-GWAS catalogue overlap of top SNPs of eosinophil count polygenic risk score. B. Location and coding/non-coding analysis of eosinophil count polygenic risk score. C. SNP-GWAS catalogue overlap of top SNPs of allergy/eczema polygenic risk score. D. Location and coding/non-coding analysis of allergy/eczema polygenic risk score.
